# Perioperative Enteral Fasting Duration Scores In Double Digits: A Seven-Year Long Cross-Sectional Retrospect By A Single Institution Among Its Patients Undergoing Extracapsular Cataract Extraction With Intraocular Lens Implantation

**DOI:** 10.1101/2023.10.18.23297215

**Authors:** Deepak Gupta

## Abstract

**Background:** Perioperative fasting period can be very variable which must be quantified at institutional level so that the institutions can re-formulate their local policies while adapting to updated national and international guidelines.

**Objective:** The purpose of this retrospective study was to ascertain the total duration of perioperative fasting among patients who underwent extracapsular cataract extraction with intraocular lens implantation at the institution during 2016-2022 (a seven-year period).

**Methods:** A query was run by information technology team in the institutional electronic medical records to extract lists of de-identified patients who underwent extracapsular cataract extraction with intraocular lens implantation at the institution during 2016-2022 wherein total perioperative fasting time was calculated from Last Food Intake to In Phase I Time (for solids) and Last Fluid Intake to In Phase I Time (for liquids).

**Results:** Data of 6722 patients was analyzed wherein patients had 4-48 hours of nothing by mouth in terms of fluid perioperatively and 10-48 hours nothing by mouth in terms of food perioperatively. Perioperative fasting durations were in double digits across all age and sex based groups whether it was the mean duration with its confidence limits or median duration with its lower and upper quartiles.

**Conclusion:** Rather than remaining in single digits, perioperative enteral fasting durations were very commonly in double digits at the institution among its patients undergoing extracapsular cataract extraction with intraocular lens implantation.

## Introduction

Preoperative fasting guidelines by the American Society of Anesthesiologists are evolving [1-2]. However, the ground reality may have remained unchanged [3]. Despite guidelines, patients are fasting for longer periods than they are required to so as to avoid delays and cancelations of their surgeries on the day of surgery. Sometimes, perioperative fasting period gets prolonged inadvertently too because of unpredictable changes in duration of other patients’ surgeries. Moreover, perioperative intravenous fluids may not be able to neutralize the effects of perioperative enteral fasting just like enteral fasting patients in medical wards [4]. The bottom-line is that perioperative fasting period can be very variable which must be quantified at institutional level so that the institutions can re-formulate their local policies while adapting to updated national and international guidelines. As there is data for long perioperative fasting periods among children [5-10], it may be time to investigate how long perioperative fasting periods are for adults undergoing common outpatient surgeries.

The purpose of this retrospective study was to ascertain the total duration of perioperative fasting among patients who underwent extracapsular cataract extraction with intraocular lens implantation at the author’s institution during 2016-2022 (a seven-year period).

## Methods

After Institutional Review Board approval for exempt research, a query was run by institution’s information technology team in the institutional electronic medical records to extract lists of de-identified patients who underwent extracapsular cataract extraction with intraocular lens implantation at the institution during 2016-2022. In those extracted lists of de-identified patients, only following data were tabulated in the query: (a) Date of Surgery and Age on the Date of Surgery, (b) their Sex/Gender, (c) Patient Arrival Time from PreOp Nursing Record, (d) Pt. In Room Time and Pt. Out Room Time from OR Nursing Record, (e) In Phase I Time from PACU I Nursing Record, and (f) Last Food Intake (Date and Time) and Last Fluid Intake (Date and Time) from Preprocedure Assessment – Adult Form’s Checklist - POH / Preprocedure. Thereafter, only total perioperative fasting time was calculated from Last Food Intake to In Phase I Time (for solids) and Last Fluid Intake to In Phase I Time (for liquids) because, having traditionally received topical anesthesia or conscious sedation for extracapsular cataract extraction with intraocular lens implantation, patients have been routinely offered food and fluid to ingest and intake orally immediately after their arrival to post-anesthesia care unit at the author’s institution.

## Results

As detailed in the CONSORT diagram (Figure 1), 1701 patients’ data out of 8423 total patients’ data got excluded most commonly due to incorrectly recorded data in the charts in terms of Last Food Intake (Date and Time) and Last Fluid Intake (Date and Time). Thereafter, only 6722 patients’ data was analyzed wherein patients had 4-48 hours of nothing by mouth in terms of fluid perioperatively and 10-48 hours nothing by mouth in terms of food perioperatively. This was under the assumptions that patients should have had a minimum of four hours perioperative fasting in terms of fluid and a minimum of ten hours perioperative fasting in terms of food. It was assumed that a minimum of two hours would have elapsed between their entries at preoperative holding areas to their entries at post-anesthesia care units while taking into account the routine time required to prepare for and perform extracapsular cataract extraction with intraocular lens implantation at the author’s institution. As detailed in Table 1, perioperative fasting durations were in double digits across all age and sex based groups whether it was the mean duration with its confidence limits or median duration with its lower and upper quartiles.

**Table 1:** Nothing By Mouth Data Among Patients Undergoing Extracapsular Cataract Extraction With Intraocular Lens Implantation During 2016-2022 Period.

| Age and sex based group of patients | Number of patients (n) | Mean ±Standard deviation for duration (hours) | 95% CI for mean duration [lower, upper] (hours) | Range for duration (hours) | Median for duration (hours) | Interquartile range for duration [lower-upper] (hours) |
| --- | --- | --- | --- | --- | --- | --- |
| **Nothing by mouth in terms of fluid perioperatively** |  |  |  |  |  |  |
| 18-64 year-old males | 1104 | 13.06 ±5.30 | [12.75, 13.37] | 4.00-40.98 | 13.07 | [10.43-15.65] |
| 18-64 year-old females | 1495 | 12.77 ±4.74 | [12.53, 13.01] | 4.02-39.32 | 12.97 | [10.58-15.43] |
| 65-89 year-old males | 1506 | 13.13 ±5.38 | [12.86, 13.40] | 4.00-43.47 | 13.07 | [10.50-15.83] |
| 65-89 year-old females | 2533 | 12.87 ±4.78 | [12.68, 13.05] | 4.00-40.80 | 12.98 | [10.48-15.65] |
| 90 or above year-old males or females | 84 | 14.16 ±5.88 | [12.91, 15.42] | 4.05-41.60 | 14.64 | [11.31-17.89] |
| **Nothing by mouth in terms of food perioperatively** |  |  |  |  |  |  |
| 18-64 year-old males | 1104 | 15.29 ±3.72 | [15.07, 15.51] | 10.00-40.15 | 14.88 | [12.70-17.08] |
| 18-64 year-old females | 1495 | 15.06 ±3.44 | [14.88, 15.23] | 10.00-44.27 | 14.60 | [12.75-16.68] |
| 65-89 year-old males | 1506 | 15.39 ±3.84 | [15.19, 15.58] | 10.00-43.47 | 14.75 | [12.69-17.22] |
| 65-89 year-old females | 2533 | 15.29 ±3.49 | [15.16, 15.43] | 10.00-42.70 | 14.88 | [12.80-17.08] |
| 90 or above year-old males or females | 84 | 16.82 ±4.92 | [15.77, 17.87] | 10.08-41.60 | 15.99 | [14.02-18.74] |

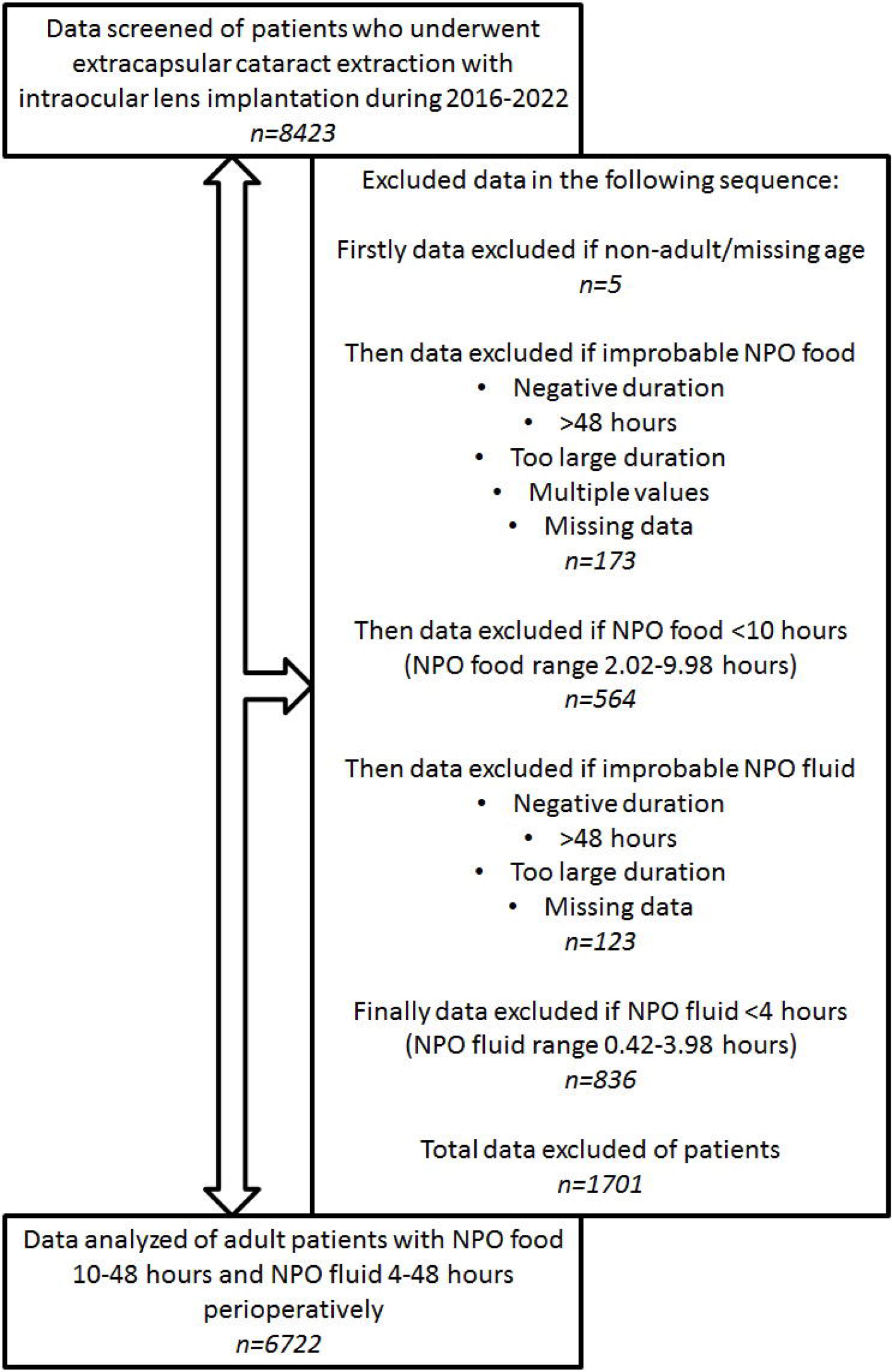

## Discussion

The key findings of this study were that (a) patients undergoing extracapsular cataract extraction with intraocular lens implantation were fasting for way longer durations perioperatively even in terms of food, and (b) these long durations of perioperative fasting in terms of fluid or food were present across the board, even among 90 or above year-old patients. These findings are similar to the findings by Falconer et al [11], who had investigated their 292 patients over a 3-month period and reported median fasting duration of >9 hours for fluids and >13 hours for food before elective surgeries and ∼13 hours for fluids and >17 hours for food before emergency surgeries. It may be specifically concerning for the practice of anesthesia if long durations of nothing by mouth for extracapsular cataract extraction with intraocular lens implantation may drive ophthalmologists and their patients to exclude anesthesia providers from their perioperative care [12], unless nothing by mouth may be warranted even by ophthalmologists in the case of topical and regional anesthesia ineffectively blocking ocular reflexes thus risking perioperative nausea and vomiting on full stomach [13-15].

There are few limitations of this study. As it was retrospective study among de-identified patients, it was not determined whether their surgeries were elective or emergent. Moreover, a large number of patients’ data got excluded due to missing entries and/or potentially erroneous entries into Last Food Intake (Date and Time) and Last Fluid Intake (Date and Time) data-fields during the completions of Preprocedure Assessment – Adult Form’s Checklist - POH / Preprocedure. Additionally, patients’ data reflecting nothing by mouth in terms of food or fluid perioperatively for >24 hours but <48 hours was also included in analysis assuming that such data might have been reflective of inpatients fasting for way longer periods due to various other reasons. Still, patients’ data analysis could have become better with exclusion of those presumed inpatients’ data to focus only on patients who had 4-24 hours of nothing by mouth in terms of fluid perioperatively and 10-24 hours nothing by mouth in terms of food perioperatively.

## Conclusion

Summarily, rather than remaining in single digits, perioperative enteral fasting durations were very commonly in double digits at the author’s institution among its patients undergoing extracapsular cataract extraction with intraocular lens implantation.

## Supporting information

IRB APPROVAL

## Data Availability

All data produced in the present work are contained in the manuscript.

