## Supplementary material for "Perioperative Enteral Fasting Duration Scores In Double Digits: A Seven-Year Long Cross-Sectional Retrospect By A Single Institution Among Its Patients Undergoing Extracapsular Cataract Extraction With Intraocular Lens Implantation": IRB APPROVAL

WAYNE STATE UNIVERSITY

IRB Administration Office
87 East Canfield, Second Floor
Detroit, MI 48201
www.irb.wayne.edu

### CONCURRENCE OF EXEMPTION
#### IRB-23-02-5543-M1 Expedited/Exempt-EXEMPT

| | |
|---|---|
| **DATE:** | May 19, 2023 |
| **TO:** | Gupta, Deepak, Anesthesiology<br>Zestos, Maria, Deans Office Medicine |
| **FROM:** | Cushing, Barbara, M1 Expedited/Exempt |
| **PROTOCOL TITLE:** | How Long Are Our Patients Fasting Perioperatively When Presenting For Extracapsular Cataract Extraction With Intraocular Lens Implantation? |
| **FUNDING SOURCE:** | None |
| **PROTOCOL NUMBER:** | IRB-23-02-5543<br>Approval Date: May 18, 2023 |

The above-referenced protocol has been reviewed and found to qualify for Exemption according to category 4

**Note to PI:** This IRB approval does not replace administrative or department/college/division approvals that may be required. Before initiating research activities contact the Associate/Vice Dean for Research in your school or college for established parameters for site access to the facility where the study will be conducted.

The following attachments and consent/assent documents have been reviewed and approved by the IRB.

Notes:

NOTE TO PI: This project has been given a Status Check-In Date. The Status Check-In Date is 05/17/2026. The Minimal Risk Status Update Form should be used to provide a status report to the IRB. Please submit the status update at least 6 weeks before this date. The Minimal Risk Status Update Form is available on the IRB's Forms and Submissions website (www.irb.wayne.edu). The Minimal Risk Status Update should be submitted as an expedited amendment via eProtocol with the Minimal Risk Status Update Form. Include the Minimal Risk Status Update Form as an Attachment using the label: Minimal Risk Status Update.

Note, if modifications to research activities and or study documents are needed; review and approval by the IRB is required BEFORE implementation. Modifications must be submitted to the IRB as an amendment.

For instructions and information on the Minimal Risk Status Report submission process see the WSU IRB ’s Guidance Tool.

Protocol (received 05/16/2023)

The following data collection materials have been reviewed and approved and does not require a WSU IRB stamp for use. These documents are approved and noted in the IRB file: (1) Data Worksheet

A waiver of HIPAA Authorization to screen medical records has been granted in accordance with the Privacy Rule and justification provided by the Principal Investigator for the HIPAA Summary Form. This waiver satisfies: 1) the use or disclosure of PHI involves no more than minimal risk to the privacy of individuals, 2) the research could not be practicably conducted without the waiver, 3) the research could not be practicably conducted without access and use of the PHI, 4) adequate steps taken to protect identifiers from improper use or disclosure and 5) adequate plan for destroying identifiers or links.

A waiver of consent has been granted according to 45CFR46.116(d). This waiver satisfies: 1) risk is no more than minimal, 2) the waiver does not adversely affect the rights and welfare of research participants, 3) the research could not be practicably carried out

e-protocol

IRB Administration Office
87 East Canfield, Second Floor
Detroit, MI 48201
www.irb.wayne.edu

without the waiver, and 4) the participant will be given information.

* Exempt protocols do not require annual review by the IRB, however you may have been granted a Status Check-In Date. Projects granted a Status Check-In date must submit a Minimal Risk Status Update Report at least 6 weeks before the check-In date. If research activities are complete a Final Report/Closure must be submitted by the Status Check-In date.

* All changes or amendments to the above-referenced protocol require review and approval by the IRB BEFORE implementation.

* Adverse Reactions/Unanticipated Problems AR/UP must be submitted on the appropriate form within the time frame specified in the IRB. In the event of an unanticipated problem use the Unanticipated Problem Report Form located on the IRB's Forms and Submissions Requirements website.

Note: Studies conducted at DMC sites or DMC medical record used for affiliate review Authorized DMC personnel have been added to this submission under Personnel Information "Other".
Administration Office Policy www.irb.wayne.edu/policies-human-research

NOTE: Upon notification of an impending regulatory site visit, hold notification, and/or external audit the IRB Administration Office must be contacted immediately. Also Notify the IRB of any changes to the funding status of the above-referenced protocol.

-------------------------------------------------------------------------------------------------------------------------------------------

**Review Type:** EXEMPT
**IRB Number:** M1 Expedited/Exempt Review

e-protocol
